# MDMA-Assisted Therapy Randomized Controlled Trial Incremental Effects Systematic Review and Meta-Analysis

**DOI:** 10.64898/2026.05.05.26352468

**Authors:** Nicholas C. Borgogna, D. Drew Whittington, Tyler Owen, Dan Petrovitch, Jacob Vaughn, Cara Struble, Louis A. Pagano, Stephen L. Aita, Benjamin D. Hill

**Author notes:** **Corresponding Author** is Nicholas C. Borgogna, University of Alabama at Birmingham, Campbell Hall, Rm 415, 1300 University Blvd. Birmingham, AL 35233.

## Abstract

Mental illness poses a substantial global burden, yet existing psychotherapies and psychopharmacologies often produce limited outcomes. Psychedelic-assisted therapies have re-emerged as potential transdiagnostic interventions. In particular, 3,4-methylenedioxymethamphetamine–assisted therapy (MDMA-AT) has generated interest for its rapid psychological effects and potential to enhance psychotherapy outcomes. However, the incremental efficacy of MDMA-AT relative to control interventions across transdiagnostic outcomes remains unclear. Further, there have been emerging concerns regarding harm reporting quality in MDMA-AT clinical trials. We conducted a systematic review and meta-analysis of MDMA-AT randomized controlled trials. Eleven publications representing eight controlled trials with 10 analyzed subgroups (*n* = 295 participants) were included in meta-analyses. Two additional secondary publications were included for harm reporting syntheses (*k* = 13 total). Across 114 extracted effect sizes, MDMA-AT demonstrated a significant moderate-to-large incremental reduction in psychopathology relative to controls (g = 1.03, 95% CI [0.46, 1.60]), though heterogeneity was high (I² = 76%). Incremental effects were larger versus inert placebos (g = 1.27) than active controls (g = 0.75). Symptom-specific analyses indicated strong incremental effects for trauma reduction (g=1.46 [95% CI: 0.67, 2.25]) and smaller non-significant effects for depression (g=0.51 [95% CI: −0.06, 1.08]). Harm reporting quality synthesis showed only 23% of publications met high-quality reporting standards. Overall, MDMA-AT demonstrates potential transdiagnostic efficacy, but small samples, confounding factors, and mediocre harm reporting highlight the need for larger more transparent clinical trials.

---

Mental illness is a serious health issue. Approximately, one-in-four people will be diagnosed with a psychopathology in their lifetime (National Institute of Mental Health, 2022). The global cost in United States (US) dollars is in the trillions (The White House, 2022). Despite the enormous social burden, solutions are limited. Psychotherapy and psychotropic medications are the two most developed treatment paradigms. Both programs have considerable research bodies. However, the quality, generalizability, and accuracy of extant psychiatric evidence-based interventions are heterogeneous. In part this is due to structural problems inherent within psychiatric science. For example, most mental illness classifications are not tied to specific biological markers (Deacon, 2013). This makes it difficult to accurately pair interventions with specific presentations, even if they are known to have some therapeutic effect. The lack of biological markers has led to a reliance on latent disease models of classification. This approach has been highly criticized for having diagnostic criteria that are arbitrary, heterogeneous, artificially comorbid, and poorly understood etiologically (Borgogna, Owen, & Aita, 2024; Deacon, 2013; Hofmann & Hayes, 2019; Insel et al., 2010). As such, there has been increased interest in fast acting “transdiagnostic” interventions (Dalgleish et al., 2020; Dindo et al., 2017).

## MDMA

MDMA (±3,4-methylenedioxymethamphetamine) is a psychoactive substance receiving increased interest as a potential agent that could be used in conjunction with psychotherapy (“MDMA-AT”) to incrementally improve transdiagnostic outcomes where other interventions have fallen short (Parrott, 2007; Smith et al., 2022). MDMA is an entactogen that acts on multiple neurotransmitter systems, with agonist effects on 5-HT1a, 2a, 2b, D1/D2, and α2 receptors (among others) leading to rapid and profound social, sensory, anxiolytic, cardiovascular and other psychological and physiological effects. Utilization of these subjective effects towards pro-social and values-driven behavior change is a key feature of MDMA-AT (Mithoefer et al., 2011) that can conceptually be applied to many conditions, including those without formal diagnostic classification (e.g., minority stress; Ching, 2020).

Experimental MDMA clinical applications date back to the 1970s, with research on precursor compounds, such as methylenedioxyamphetamine originating even earlier (Passie, 2018). However, throughout the 1980’s MDMA rose to prominence as a recreational substance tied to crime, sexual assault, and addiction. In 1985, MDMA was declared a Schedule I substance with no currently accepted medical use (Climko et al., 1986), leading to a decline in therapeutic research and increased stigmatization as a drug of abuse. While there have been significant efforts to reclassify MDMA in the US (Siegel et al., 2023), it remains a Schedule I substance.

The 2010s ushered in the so-called *psychedelic renaissance* (Rhee et al., 2023), with psychedelic agents such as psilocybin, ketamine, and many other classic and non-classic psychedelics receiving increased attention as potential transdiagnostic solutions to treatment resistant mental illness. In particular, there has been excitement that psychedelic-assisted therapy (PAT) broadly can be applied to diverse/transdiagnostic conditions and that the beneficial effects are rapid (Fadiman & Korb, 2019; Rhee et al., 2023; Schenberg, 2018). MDMA has been included in this resurgence and accompanying de-stigmatization. A specific form of MDMA-AT (midomafetamine) received “breakthrough” status as a treatment for post-traumatic stress disorder (PTSD) in 2017. However, Food and Drug Administration (FDA) approval was denied in 2024, after several design features were identified that precluded favorable review (including blinding problems, lack of long-term follow-up, unreported adverse events, participant selection bias, and protocol violations). FDA denial was further followed by the redaction of three prominent MDMA publications (Feduccia et al., 2021; Jerome et al., 2020; Mithoefer et al., 2019), and an ensuing ethics controversy (patient sexually assaulted by MDMA-AT staff during phase II clinical trial; Harrison et al., 2025).

Notwithstanding these shortcomings, PAT broadly and MDMA-AT specifically provide a potential framework for moving beyond the extant stasis in mental illness intervention. Psychedelics are generally associated with rapid acting effects, addressing a key problem in current psychotherapy and medication treatments. Further, they potentially offer an opportunity for intensive, but short term, therapy programs. That is, they potentially reduce the costs and burden of having to take daily medication, such as side effects and treatment withdrawal. Not surprisingly, positive results from initial clinical trials from across the PAT spectrum have been associated with considerable public interest (Abdallah et al., 2022; Lamotte, 2022; Mitchell et al., 2023; von Rotz et al., 2023).

However, the incremental efficacy of this potential has yet to be fully demonstrated. While many trials have demonstrated symptom reductions following psychedelic agent administration, the degree to which the effect is superior to competing evidence-based agents is often limited. For example, multiple meta-analyses have demonstrated that ketamine reduces PTSD symptoms, but that the size of the reduction is small compared to convincing control agents that have similar physiological effects (Borgogna, Owen, Vaughn, et al., 2024; Sicignano et al., 2024). This was recently demonstrated in a clinical trial comparing ketamine infusion to midazolam in the treatment of depression (Jelovac et al., 2025). Therapeutic effects were observed, but not incrementally superior to midazolam. Putatively, such results demonstrate PAT benefits are due to placebo/expectancy effects and/or variations in the psychotherapy.

Our position is that if MDMA-AT is to continue being considered as a potential paradigm shifting intervention it should be able to withstand critical incremental efficacy tests. In other words, how much better is MDMA-AT at reducing transdiagnostic mental illness symptoms relative to control interventions, which outcomes demonstrate the strongest incremental reduction, and is the effect stronger against certain types of controls? While multiple meta-analyses have demonstrated MDMA-AT is associated with mood enhancements and symptom reductions (Amoroso & Workman, 2016; Shahrour et al., 2024; Smith et al., 2022), few quantified transdiagnostic incremental effects relative to control conditions. That is, as far as we are aware, all MDMA-AT RCTs include multiple symptom outcome measures, yet meta-analyses tend to target one diagnostic outcome (e.g., PTSD symptoms). For example, Mithoefer et al. (2018) targeted PTSD, but also included six additional secondary assessments. Simply, examining a single outcome for a review misses the bigger picture, especially as the field moves towards transdiagnostic approaches. Thus, it would be important to understand omnibus meta-analytic effects across measures. However, it is also important to compare standardized effects in relation to different types of controls. Cumulatively, such an analysis will help researchers, clinicians, and stakeholders understand how much gain might be associated with MDMA-AT as a transdiagnostic intervention.

Concurrently, it is important to quantify the risks associated with MDMA-AT. Considerable work has demonstrated that MDMA is addictive (Pantoni et al., 2022) and potentially fatal (Armenian & Rodda, 2022; Schifano, 2004). To be sure, MDMA-AT involves considerably more structure and is putatively safer than recreational MDMA use. However, Colcott and colleagues (2025) recently published a review and meta-analysis of side-effects and risk of bias in MDMA-AT clinical trials. They reported almost all of the reviewed MDMA-AT trials as having a high harm reporting bias risk. Further, none of the studies they reviewed could pass more than 64% of the CONSORT Harms 2022 guidelines for harm reporting quality – suggesting potential harms are not being reporting or not being reported in convincing detail. However, Colcott et al. (2025) only reviewed eight studies for Harm Reporting Quality (HRQ). Moreover, not all of the studies were randomized controlled trials (RCTs). Further, several of the published MDMA-AT RCTs are associated with secondary and exploratory papers that retain the original controlled trial framework (e.g., Gorman et al., 2020; Ponte et al., 2021), but are typically excluded from reviews. While these publications typically reference their parent publication, they also pose a potential bias risk by reporting preferable outcomes (e.g., symptom reductions) without full harm data, which might be described in the parent paper, but not the secondary analysis. It is unclear the degree to which secondary MDMA-AT report harms from their parent trials. We sought to build upon the identified gaps by conducting a systematic review and meta-analysis with the following specific aims:

1. Conduct an incremental effects meta-analysis of all MDMA-AT RCTs.

a. Test the overall incremental effect where all outcome measures are examined.
b. Test overall incremental effects of MDMA-AT compared to active controls (psychoactive agents) and inert placebos.
c. Test incremental effects of MDMA-AT for specific mental illness outcomes. This sub-aim is important because many MDMA-AT clinical trials include target specific diagnostic profiles (e.g., PTSD) but include outcome measures from several different symptom profiles.
2. Synthesize HRQ findings by reviewing HRQ data from all MDMA-AT publications that retain a controlled trial/RCT framework, including secondary publications.

## Methods

### Search Strategy

The Preferred Reporting Items for Systematic Reviews and Meta-Analyses (PRISMA) guidelines informed this study. Covidence software was used to manage searches, screenings, and extractions. A medical librarian was consulted to develop standardized search terms for the following databases: PubMed, Embase, Health & Medical Collection, PsycINFO, and Psychology Database. Search terms were: MDMA OR methylenedioxymethamphetamine OR Ecstasy AND (PTSD OR Post-Traumatic OR Posttraumatic OR Depress* OR MDD OR Mood OR Affective OR Anxiety OR Anxious OR Panic OR Fear OR OCD OR Obsessive-Compulsive Disorder OR Scrupulosity OR Obsess* OR Compuls* AND trial* OR study). We also compared our identified RCTs to published MDMA-AT meta-analyses to ensure no RCTs were missed.

### Inclusion/Exclusion Criteria

The following inclusion criteria were utilized: a) Human subjects, b) Employment of continuously scaled mental illness outcome measures c) Controlled trial design, d) MDMA-AT experimental condition. Exclusion criteria were a) Non-peer review publication outlet, b) Dosage studies without controls (e.g., control subjects exposed to MDMA macro doses – MDMA micro dose control conditions were considered acceptable), c) MDMA applied for severe medical/non-psychiatric issues, d) Pediatric or adolescent samples (under the age of 18), and e) Secondary analyses of original RCT data were considered exclusionary for aim 1. English language publication was *not* an exclusion criterion. Cross-over trials were accepted if they provided outcomes prior to cross-over.

### Data Extraction

After completing the initial literature search, titles and abstracts were assessed. If the abstract suggested the study could meet inclusion criteria, the full text was reviewed by at least two study team members. Studies that met inclusion criteria had means (mean change when raw means were not available), standard deviations/errors, *n*’s, and basic demographic information extracted for analyses. Any extraction/coding issues were resolved via team discussion.

Figure 1 illustrates our literature search. In total, *k*=1,526 papers were identified with 1,493 screened after deduplication, and 13 identified for analyses. However, two of these papers (Ponte et al. 2021; Gorman et al. 2020) were secondary analyses that pooled outcomes from previously published RCTs. To maintain statistical independence, estimates from these papers were not included in meta-analyses. Further, three studies (Brewerton et al., 2022; Nicholas et al., 2022; van der Kolk et al., 2024) were part of parent RCT (Mitchell et al., 2021). Distinct outcomes from these three studies were nested within the parent RCT for meta-analyses, but no duplicate measures were included. Two studies included two active treatment conditions compared to a control (Mithoefer et al., 2018; Ot’alora G et al., 2018). In this case, active conditions were treated as separate subgroups against controlled comparisons, but active arms were not compared to one another. In total, eleven papers (Brewerton et al., 2022; Danforth et al., 2018; Mitchell et al., 2021, 2023; Mithoefer et al., 2011, 2018; Nicholas et al., 2022; Oehen et al., 2013; Ot’alora G et al., 2018; van der Kolk et al., 2024; Wolfson et al., 2020), representing eight trials, with 10 analyzed subgroups were included for aim 1. All 13 papers were synthesized for aim 2 evaluating HRQ (including Gorman et al., 2020 and Ponte et al., 2021).

**Figure 1.**
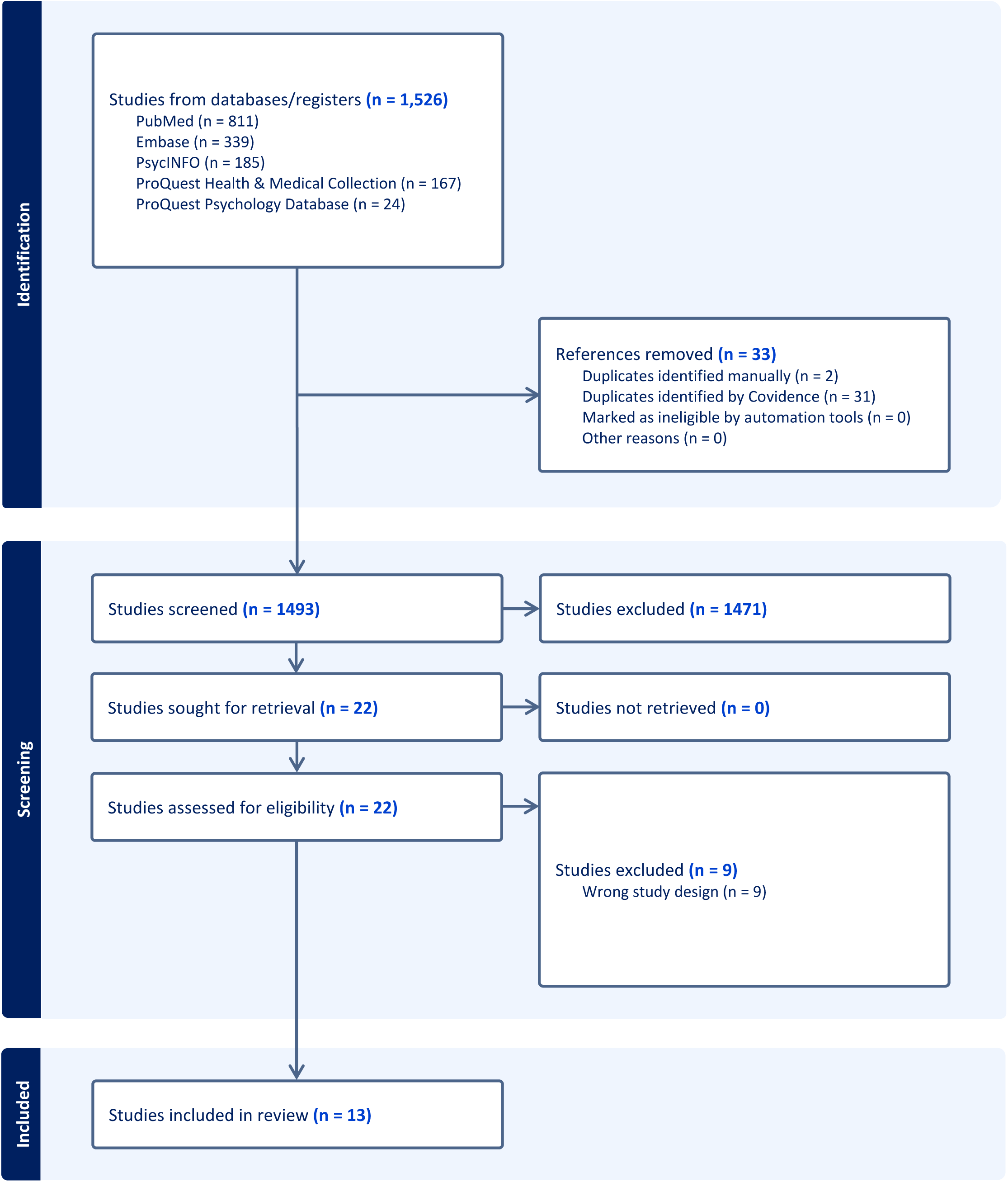
Search Outcomes.

If we could not obtain specific estimates, we extracted estimates from figures using the Web Plot Digitizer tool (version 4) from Automeris (2024). Mitchell et al. (2023) was the only study that neither reported pre/post means or standard deviations for outcomes nor responded to author requests for estimates. Mitchell et al. (2021) also did not report pre/post means and standard deviations, but provided specific within group effect sizes and 95% confidence intervals that were used to estimate incremental effects.

### Harm Reporting Quality

The CONSORT Harms 2022 checklist (Junqueira et al., 2023) guided our synthesis. Questions were evaluated on a binary scale with a score of “1” denoting satisfied criteria. All papers were scored by at least two reviewers. Discrepancies were resolved by involving a third reviewer and, when needed, group discussion. Only original manuscript materials were evaluated during HRQ synthesis. Statements linking materials to supplementary files or external links were considered unsatisfactory. The CONSORT checklist involves many barreled criteria. All elements of a criteria needed to be met to receive a satisfactory “1” score. Scores for each item on the checklist were summed and categorized as high quality (meeting at least 80% of the checklist); moderate quality (meeting between 50-80% of the checklist); or low quality (meeting less than 50% of the checklist).

### Incremental Efficacy Data Analysis

Effect sizes were analyzed using Comprehensive Meta-Analysis (CMA v4) software (Borenstein et al., 2013). All analyses were modeled under random effects. Hedges’ g was selected as the index of effect size. Post-score standard deviation was used to standardize the effect sizes given pre-post correlations were generally not reported. In all analyses, we examined the incremental effect size of MDMA-AT experimental intervention relative to control. That is, we compared the within group change across timepoints between conditions. Incremental Hedges’ g represents the strength of the treatment group (MDMA-AT) above-and-beyond the control group in reducing mental illness and/or improving life quality. Consistent with past meta-analyses (Borgogna et al., 2025; Pizer et al., 2024), we adopted Ferguson’s (2009) recommendation of Hedges’ g ≥ 0.41 as the threshold for minimal “practical” significance.

Heterogeneity was estimated using τ, Q, and I^2^ values. Indicators were coded such that positive values represented a greater therapeutic effect in the MDMA-AT conditions relative to controls. When studies employed multiple measures, standardized means between the measures were estimated. For example, if a study employed the Beck Depression Inventory-II [BDI-II] and Montgomery–Åsberg Depression Rating Scale [MADRS], standardized effects from both measures were estimated and then pooled. Across analyses, study subgroups were set as the unit of analysis. Publication bias was meta-analytically assessed using Egger’s regression test (Egger et al., 1997). Duval and Tweedie’s trim-and-fill method was used to assess the possibility of missing studies due to publication bias (Duval & Tweedie, 2000). Each nested measure was adjusted by reported *n*. No missing data was estimated.

For aim 1a, we examined all reported outcomes comparing MDMA-AT to controls. To test aim 1b, we conducted sub analyses examining the incremental effects between active and passive control conditions. To test aim 1c, we conducted a series of subgroup analyses for specific symptoms: PTSD (Clinician Administered PTSD Scale, Impact of Event Scale-Revised, and Posttraumatic Diagnostic Scale), depression (BDI-II and MADRS), and “other” psychopathology measures (Alcohol Use Disorder Identification Test, Dissociative Experiences Scale-II, Drug Use Disorders Identification Test, Eating Attitudes Test-26, Emotion Regulation Questionnaire, Interpersonal Reactivity Index [personal distress subscale], Leibowitz Social Anxiety Scale, Neuroticism-Extroversion-Openness-Personality Inventory-Revised [neuroticism subscale], Perceived Stress Scale, Pittsburgh Sleep Quality Index, State-Trait Anxiety Inventory, and Toronto Alexithymia Scale-20). See Supplemental File 1 for measure references. To address aim 2, HRQ data were summed for each study. Because the nature of aim 2 is descriptive, results from HRQ syntheses are reported first, followed by formal inferential meta-analyses.

### Registration

Aims were developed in July 2024. However, FDA denial of MDMA-AT (August 2024) was associated with study reconfigurations. Initial literature searches began in August 2024. Formal registration occurred in 2025 (https://osf.io/jbrny) after an initial search had been completed, but prior to analyses. Follow-up ad-hoc literature searches, post-registration, failed to identify any additional studies as of March 2026.

## Results

In total, *n*=295 (*n*=172 MDMA experimental condition assignment) participants were analyzed across the included studies. Table 1 describes basic demographic features. With exception to Mithoefer et al. (2018) and Danforth et al. (2018), all of the studies involved samples primarily comprised of women. Danforth et al.’s (2018) was also unique in that involved autistic participants and had a control group entirely comprised of men. Across studies 17-56% of participants had prior MDMA and/or other psychedelic experience (Ot’alora G et al. did not report). MDMA-AT experimental dosing approaches varied across studies, ranging from 75 mgs (Mithoefer et al. 2018) to 180 mgs (Mitchell et al. 2021). Danforth et al. (2018) had two

**Table 1.** Demographics.

Demographics
| Reference | %Women | %Non-Naïve | #Dosing Sessions | #Study Sites | %Medication Taper |
| --- | --- | --- | --- | --- | --- |
| Oehen et al. (2013) | 83% | 17% | 3 | 1 | 50% |
| Wolfson et al. (2020) | 77.8% | 55.6% | 2 | 1 | 33.3% |
| Mithoefer et al. (2018) | 27% | 23% | 2 | 1 | * |
| Ot'Alora G et al. (2018) | 67.9% | NR | 2 | 1 | ** |
| Mitchell et al. (2021) | 65.6% | 32.2% | 3 | 15 | 25.5% |
| Mithoefer et al. (2011) | 85% | 45% | 2 | 1 | ~75%*** |
| Mitchell et al. (2023) | 71.15% | 46.15% | 3 | 13 | 26% |
| Danforth et al. (2018) | 16.67%**** | 0% | 2 | 1 | ***** |

Note: Non-Naïve, refers to participants who reported histories of recreational MDMA/psychedelic use.
Medication taper refers to the number of participants required to taper/washout current psychiatric medication prior MDMA-AT. NR=Not reported.
\* Mithoefer et al. (2018) reported number of pre-study medications, but not by participant. Across participants there was a history of 89 prescriptions (for 26 participants). Their appendix indicated four participants took psychiatric medications during follow-up.
\*\*Ot'Alora G et al. (2018) reported number of pre-study medications, but not by participant. Across participants there was a history of 58 prescriptions (for 28 participants). Based on their supplementary file, at least 10 participants resumed medication after the study all of which were reported to have prior medication histories.
\*\*\*Mithoefer et al. (2011) reported "Fifteen of the 20 subjects had previously undergone multiple medication trials (mean 4.2 different psychiatric drugs)" (p.445).
\*\*\*\*Danforth et al. (2018) had a control condition exclusively comprised of men. A participant was also removed after baseline in their MDMA-AT condition, but their gender was not reported.
\*\*\*\*\*Danforth et al. (2018) reported number of pre-study medications, but not by participant. Across participants there was a history of 18 prescriptions (for 12 baseline participants).

MDMA-AT dosing conditions (75-100 and 100-125 mgs) but reported combined results. Exact protocols differed across studies regarding the amount, time, and structure of intervention (e.g., employment of supplemental half doses) and outcome measurements. Controls were all either inert placebos or subtherapeutic (25-40 mgs) MDMA doses (Mithoefer et al. 2018; Ot’alora G et al., 2018; Oehen et al. 2013). Most studies involved a washout period in which participants were required to discontinue any current psychiatric medications prior to MDMA-AT. Of note, all studies received support from the Multidisciplinary Association for Psychedelic Studies (MAPS).

### Harm Reporting Quality

HRQ results are available on Table 2. The formal CONSORT criteria used in evaluation are available in Supplemental File 2. Only 23% (*k*=3) of the papers were considered to have “high quality” reporting, with six having moderate quality, and four having low quality. Notably, all four “low quality” papers were secondary analyses. Ot’alora G et al. (2018) was considered to be the best quality meeting >90% of the criteria, whereas Gorman et al. (2020) was considered the worst (<40%). Several HRQ items were met by all studies (e.g., criteria 18*: Results of any other analyses performed, including subgroup analyses and adjusted analyses, distinguishing pre-specified from exploratory*). Criteria 17a (*For each primary and secondary outcome of benefits and harms, results for each group, and the estimated effect size and its precision (such as 95% confidence interval)*) and 17b (*Presentation of both absolute and relative effect sizes is recommended, for outcomes of benefits and harms*) were the most challenging and not fully met by any paper. In large part, this has to do with the barreled nature of the items focusing on effect sizes/precision estimates across both benefits and harms. Most articles met at least part of the criteria (generally as it relates to benefits). Given the nature of HRQ, we opted for conservative interpretations.

**Table 2.**
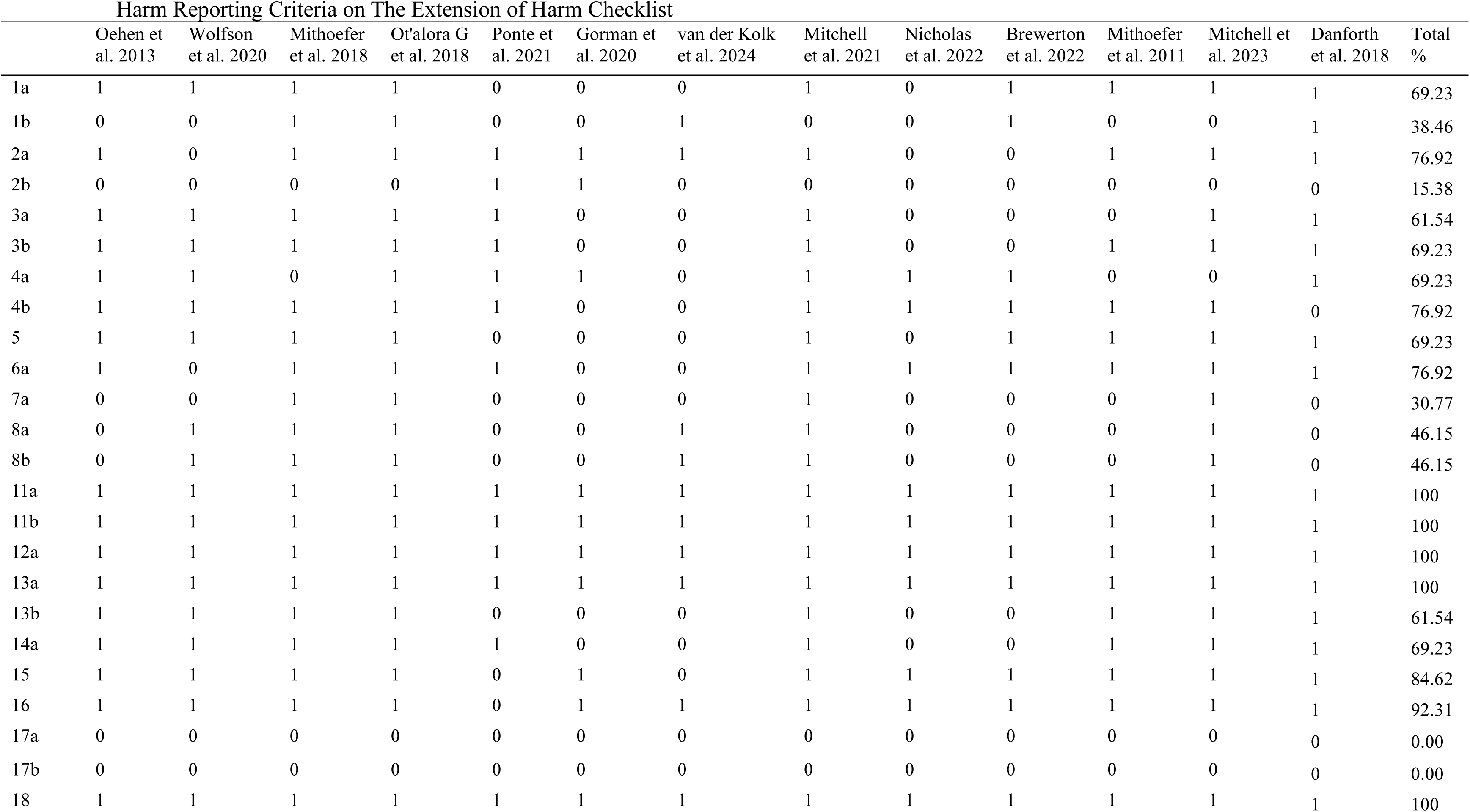
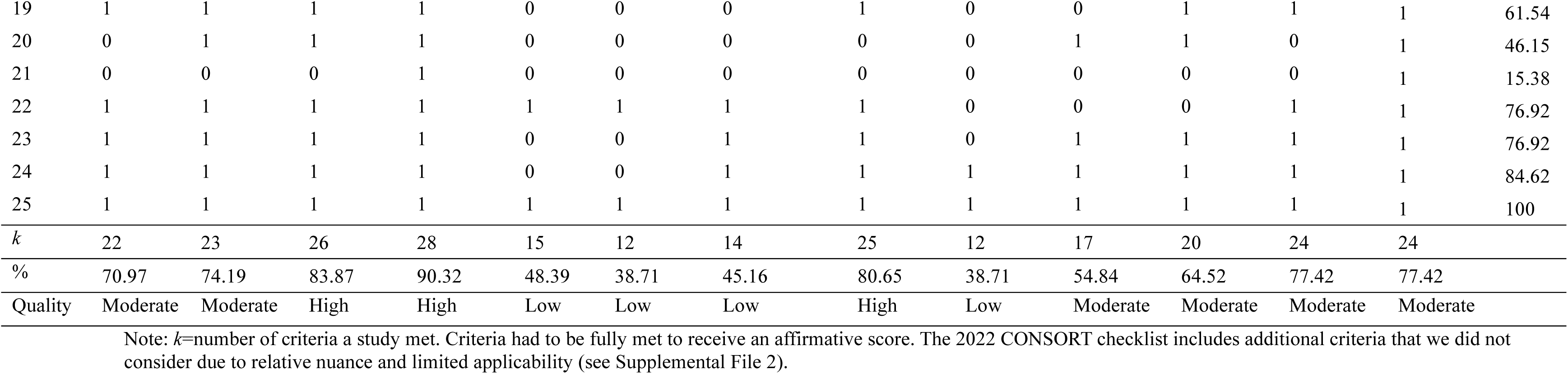
Harm Reporting Criteria on The Extension of Harm Checklis.

### Incremental Meta-Analyses

Across the included studies, 114 effect sizes were extracted and pooled using a three-level structure. Table 3 demonstrates pooled effect sizes by study. Results indicated that across time points, measures, and subgroups, MDMA-AT is associated with a moderate significant incremental effect size of g=1.03 (95% CI: 0.46, 1.60), prediction interval=-0.87, 2.93, τ=0.77, Q(9)=36.71, *p*<.001, I^2^=76% in terms of reducing psychopathology and increasing functioning relative to controls. Duval and Tweedie’s trim and fill did not suggest any adjustments, but these results should be interpreted cautiously. Moreover, the funnel plot (Supplemental File 3) had *k*=three subgroups outside the funnel with seven inside (eight to the left of the effect – meaning it is inflated). The effect is observably biased by two studies with unusually large incremental effect sizes (Mithoefer et al., 2011 [3.42] and Mitchell et al., 2023 [1.97]). Egger’s regression was non-significant, b_0_=-0.17, *p*=.35.

**Table 3.** Combined Incremental Effects by Subgroup.

Combined Incremental Effects by Subgroup
| Reference | Dose | Control | *n* |  | Hedges *g* | 95% CI | Relative Weight |
| --- | --- | --- | --- | --- | --- | --- | --- |
|  |  |  | MDMA | Control |  |  |  |
| Oehen et al. (2013) | 125 mg <sup>a</sup> | 25 mg <sup>b</sup> | 8 | 4 | 0.70 | -0.48, 1.87 | 8.90 |
| Wolfson et al. (2020) | 125 mg <sup>a</sup> | Placebo | 13 | 5 | 0.46 | -0.55, 1.46 | 9.90 |
| Mithoefer et al. (2018) | 125 mg <sup>a</sup> | 30 mg <sup>d</sup> | 11 | 6 | 1.07* | 0.05, 2.09 | 9.83 |
| Mithoefer et al. (2018) | 75 mg <sup>c</sup> | 30 mg <sup>d</sup> | 5 | 6 | 1.23 | 0.00, 2.45 | 8.64 |
| Ot'Alora et al. (2018) | 100 mg <sup>e</sup> | 40 mg <sup>f</sup> | 9 | 6 | 0.61 | -0.41, 1.63 | 9.83 |
| Ot'Alora et al. (2018) | 125 mg <sup>a</sup> | 40 mg <sup>f</sup> | 12 | 6 | 0.36 | -0.58, 1.30 | 10.28 |
| Mitchell et al. (2021) | 80-180 mg <sup>g</sup> | Placebo | 42 | 40 | 0.45 | -0.07, 0.97 | 12.78 |
| Mithoefer et al. (2011) | 125 mg <sup>a</sup> | Placebo | 12 | 8 | 3.42** | 2.05, 4.79 | 7.86 |
| Mitchell et al. (2023) | 80-180 mg <sup>g</sup> | Placebo | 53 | 50 | 1.97** | 1.49, 2.45 | 13.00 |
| Danforth et al. (2018) | 75-125 mg <sup>f</sup> | Placebo | 7 | 4 | 0.33 | -0.83, 1.49 | 8.99 |
| Omnibus Effect | - | - | 172 | 123 | 1.03*** | 0.46, 1.60 | 100 |

Note: All outcome measures have been combined. Results from van der Kolk et al., 2024; Brewerton et al. 2022; and Nicholas et al., 2022 are nested within Mitchell et al. 2021. Occasional attrition occurred for specific outcomes (e.g., Dissociative Experiences Scale-II in Mithoefer et al. 2018), analyses have been adjusted for author reported attrition on specific outcome measures. \* $p < .05$ , \*\* $p < .001$ .
<sup>a</sup> Signifies the inclusion of a 62.5 mg supplemental dose
<sup>b</sup> Signifies the inclusion of a 12.5 mg supplemental dose
<sup>c</sup> Signifies the inclusion of a 37.5 mg supplemental dose
<sup>d</sup> Signifies the inclusion of a 15 mg supplemental dose
<sup>e</sup> Signifies the inclusion of a 50 mg supplemental dose
<sup>f</sup> Signifies the inclusion of a 20 mg supplemental dose
<sup>g</sup> Each participant received an initial dose of 80 mg followed by supplemental 40 mg dose. Follow-up sessions included 120 mg dose followed by 60 mg supplement.
<sup>f</sup> MDMA-AT condition was split into a 75-100 mg condition and a 100-125mg condition.

For studies testing MDMA-AT against inert placebos, the incremental effect was significant g=1.27 (95% CI: 0.28, 2.26). For studies testing MDMA-AT against active controls the effect was notably weaker, though still significant g=0.75 (95% CI: 0.27, 1.23). The difference in effect size was not statistically significant (*p*=.43). Though this should be interpreted cautiously given the limited number of studies. MDMA-AT’s incremental effect on PTSD measures trended slightly stronger than the overall incremental effect, g=1.46 (95% CI: 0.67, 2.25), τ=1.02, Q(7)=39.31, *p*<.001, I^2^=82%. Depression was associated with a more modest incremental effect that was non-significant: g=0.51 (95% CI: −0.06, 1.08), τ=0.57, Q(6)=13.61, *p*=.034, I^2^=56%. The other psychopathology measures produced a modest statistically significant incremental effect: g=0.57 (95% CI: 0.26, 0.88), Q(6)=3.70, *p*<.72.

## Discussion

Our systematic review and meta-analyses indicate MDMA-AT is associated with a significant incremental reduction in psychopathology outcomes relative to controls. Across all measures, MDMA-AT improved symptoms by a metric of about one standard deviation above-and-beyond controls. However, the effect was heterogenous, indeed partly due to the aggregation of diverse psychopathology measures. The effect was observably stronger when the control was an inert placebo but was still significant compared to low-dose MDMA active controls. The effect trended stronger for PTSD outcome measures, which makes sense given most trials targeted PTSD. All incremental effects were above our *a priori* threshold for practical meaningfulness (0.41), suggesting real-world clinical significance. This includes depression measures, though these were actually not statistically significant. In part, this is due to the control condition in Danforth et al. (2018) having superior BDI-II reductions relative to MDMA-AT. Depending on threshold guideline, all observed incremental effects would either be considered large (Cohen, 1988) or at least moderate (Ferguson, 2009).

The positive findings are qualified by more critical design concerns and HRQ. The instability of effect sizes is something that should be evaluated critically. Interestingly, only three out of ten incremental effects were statistically significant when accounting for the number of measures nested within each study, though they all trended in a MDMA-AT favorable direction. This is due to researchers including too many measures within their studies, relative to number of participants. For instance, in Danforth et al. (2018) 16 scales/subscales are employed despite having a final *n*=11. With exception to Mitchell et al. (2021, 2023), none of the studies had adequate distributions for the types of parametric statistics conducted. Fourteen of the 18 (>77%) examined conditions had samples *n*≤13. Decline effects can be expected in future studies, and to some extent, are already apparent. For example, Mithoefer et al. (2011) generated an almost unprecedented incremental g=3.42. However, the MDMA-AT group had only 12 participants, and the control only eight, which may be why the effect dramatically shrinks in Mitchell et al. (2021; *n*=82) to g=0.45.

This concern is compounded when considering control type. For instance, Oehen et al. (2013) has an even smaller sample than Mithoefer et al. (2011), but the incremental effect is g=0.70. The difference is that Oehen et al. (2013) employed an active control condition, which is why the incremental effect size is about 80% smaller. Also concerning is that the two largest studies (Mitchell et al. 2021, 2023) utilized inert placebos. It is possible that if a large sample (*n*’s >50 per condition) were gathered to test MDMA-AT against a convincing discrete active control condition, the incremental effects would probably be attenuated further. This has been demonstrated in pharmacological trials such as risperidone (Krystal et al., 2011) and ketamine for PTSD (Abdallah et al., 2022).

We also observed problems with HRQ. Our results were similar to Colcott et al. (2025). Arguably the biggest issue was the piecemeal dissemination process. Authors were able to generate multiple publications from the same data or combined data to produce new papers. This is not uncommon practice, and all papers acknowledged parent sources. However, in the context of MDMA-AT, this practice is concerning. Combined, less than 300 participants have actually been documented in MDMA-AT RCTs. In other words, there is a familywise error concern (hence, several study-level non-significant incremental effects). Further, adverse event information tended to be partially reported in original texts and often relegated to supplementary materials or external links. This creates a discrepancy, where readers might easily find positive MDMA-AT reports, but have to search more vigorously to find harm information. The general problem of downplaying and/or underreporting harms has been an ongoing issue in PAT (De Giorgi & Ede, 2024; Taillefer de Laportalière et al., 2023). We encourage MDMA-AT researchers to rigorously approach harm reporting, such as involving third-party scientists gathering harms data and better balancing studies with harm information in secondary analyses. If secondary analyses are approached on clinical trial data, they should account for familywise error, demonstrate satisfied statistical assumptions, and report harm information from the parent trial. We encourage editors to allow space for such information.

Another problem we observed was that patients received differing dosages across and within trials. The field has moved towards a preference for supplemental half dosages. However, even in the most recent RCT, inconsistencies arose (e.g., “*Within the MDMA-AT group, three participants did not undergo dose escalation in experimental sessions 2 and 3, and two participants experienced dose administration timing errors*” Mitchell et al. 2023 p. 2481). To some extent this is understandable as patients have various body-types, metabolisms, and other physiological features that necessitate dosage adjustments. However, this also highlights one of the risks associated with MDMA-AT in that it is not clear what might happen with a particular dose for a particular person.

### Limitations

Our study is limited by the literature body size. We consider our review to be pilot and subject to larger MDMA-AT clinical trials. Because the literature body is small we chose to limit the number of meta-analytic tests we performed. Many important subgroup and moderator tests would be preferable if the literature expands. We also did not assess outcomes per specific outcome time point (e.g., six months versus a year). This had to do with the heterogeneity across time points and lack of controlled follow-ups. A key prospective advantage of PAT is the idea that one or two dosing sessions might fundamentally change someone’s psychology, negating the need for ongoing treatment. It would be beneficial to have blinded studies with long term (>six months) outcome measurements.

Our syntheses were limited by the evaluation tools. HRQ was informed by the CONSORT statement which involved many items with barreled criteria. In practice, many authors met partial criteria. This problem has arisen in previous PAT meta-analyses (e.g., Borgogna et al., 2025). More liberal thresholds would have yielded more favorable ratings. Given the seriousness of psychopathology, and the degree to which MDMA-AT is being considered a paradigm-shifting intervention, we chose the most conservative synthesis approach. Future MDMA-AT researchers could improve their work by fully adhering to harm reporting CONSORT guidelines. A broader opportunity for clinical trial researchers would be to develop HRQ criteria tools that are not barreled.

### Conclusions

Altogether, we believe there is psychotherapeutic potential in MDMA-AT, but that design and reporting structures need to be improved. We anticipated some degree of financial conflict of interest to be associated with MDMA-AT. However, we were surprised that all RCTs were funded by MAPS. It would be preferable to examine MDMA-AT RCT outcomes from a research team without MAPS affiliation. We encourage federal bodies to fund MDMA-AT clinical trial research, but to teams without ideological conflicts of interest. If MDMA-AT can be shown to robustly and incrementally improve outcomes in studies designed with the highest level of rigor, then MDMA-AT has the potential to ameliorate the diverse arrays of psychopathology challenging society.

## Data Availability

All data produced in the present study are available upon reasonable request to the authors

## Acknowledgements

The authors would like to thank Breanna Roberts, Rachael Rowland, and Jason G. Smith for their assistance with this project.

The authors declare no conflicts of interest.

## Supplemental File 1 Measure References

American Psychiatric Association. (2000). Diagnostic and statistical manual of mental disorders: 4th edition. Arlington, VA: American Psychiatric Association.

Baer, R. A., Smith, G. T., Hopkins, J., Krietemeyer, J., & Toney, L. (2006). Using self-report assessment methods to explore facets of mindfulness. *Assessment*, 13, 27–45.

Bagby, R. M., Taylor, G. J., & Parker, J. D. A. (1994). The twenty-item Toronto Alexithymia Scale: II. Convergent, discriminant, and concurrent validity. *Journal of Psychosomatic Research, 38*, 33–40.

Baker, S. L., Heinrichs, N., Kim, H.-J., & Hofmann, S. G. (2002). The Liebowitz social anxiety scale as a self-report instrument: A preliminary psychometric analysis. *Behaviour Research and Therapy, 40*(6), 701–715.

Beck, A. T., Steer, R. A., & Brown, G. K. (1996). Manual for the Beck Depression Inventory-II. The Psychological Corporation.

Berman A.H., Palmstierna T., Källmén H., Bergman H. The self-report Drug Use Disorders Identification Test: Extended (DUDIT-E): reliability, validity, and motivational index. *Journal of Substance Abuse Treatment*, 32, 357-69.

Blake, D. D., Weathers, F. W., Nagy, L. M., et al. (1990). A clinician rating scale for assessing current and lifetime PTSD: the CAPS-1. *Behavior Therapy*, 13, 187–88.

Briere, J., & Runtz, M. (2002). The Inventory of Altered Self-Capacities (IASC): A standardized measure of identity, affect regulation, and relationship disturbance. *Assessment*, 9, 230-239.

Buysse, D. J., Reynolds, C. F. 3rd., Monk, T. H., Berman, S. R., & Kupfer, D. J. (1989). The Pittsburgh sleep quality index: a new instrument for psychiatric practice and research. *Psychiatry Research*, 28, 193–213.

Carlson, E. B., & Putnam, F. W. (1993). An update on the Dissociative Experiences Scale. *Dissociation*, 6, 16–27.

Cella, D., & Nowinski, C. J. (2002). Measuring quality of life in chronic illness: the functional assessment of chronic illness therapy measurement system. *Archives of Physical Medicine and Rehabilitation*, 83, S10-17.

Cohen, S., Kamarck, T., & Mermelstein, R. (1983). *Perceived Stress Scale*. APA PsycTests.

Costa, P. T., & Macrae, R. R. (1985). The NEO personality inventory manual. Psychological Assessment Resources.

Davis, M. H. (1980). A multidimensional approach to individual differences in empathy. JSAS Catalog of Selected Documents in Psychology, 10, 85

Foa, E. B., Riggs, D. S., Dancu, C. V., & Rothbaum, B. O. (1993). Reliability and validity of a brief instrument for assessing post-traumatic stress disorder. *Journal of Traumatic Stress*, 6, 459–473.

Garner, D. M., Olmsted, M. P., Bohr, Y., & Garfinkel, P. E. (1982). The eating attitudes test: Psychometric features and clinical correlates. *Psychological Medicine*, 12, 871-878

Gesser, G., Wong, P. T. P., & Reker, G. T. (1987–1988). Death attitudes across the life span: The development and validation of the Death Attitude Profile (DAP). Omega (Westport), 18, 113–128.

Gross, J.J., & John, O.P. (2003). Individual differences in two emotion regulation processes: Implications for affect, relationships, and well-being. Journal of Personality and Social Psychology, 85 , 348-362.

McDonald, S., Bornhofen, C., Shum, D., Long, E., Saunders, C., & Neulinger, K. (2006). Reliability and validity of The Awareness of Social Inference Test (TASIT): A clinical test of social perception. *Disability and Rehabilitation*, *28*, 1529–1542.

Montgomery, S. A., & Asberg, M. (1979). A new depression scale designed to be sensitive to change. *British Journal of Psychiatry*, 134, 382–389.

Neff, K. (2003). The development and validation of a scale to measure self-compassion. *Self Identity*, 2, 223–250.

Rosenberg, M. (1979). Conceiving the Self. New York: Basic Books.

Saunders J.B., Aasland O.G., Babor T.F., de la Fuente J.R., Grant M. (1993). Development of the Alcohol Use Disorders Identification Test (AUDIT): WHO Collaborative Project on Early Detection of Persons with Harmful Alcohol Consumption--II. *Addiction*, 791-804.

Schalock, R. L., & Keith, K. D. (1993). *Quality of Life Questionnaire (QOL.Q)*. APA PsycTests.

Sheehan, D. V., Harnett-Sheehan, K., & Raj, B. A. (1996). The measurement of disability. *International Clinical Psychopharmacology, 11*, 89–95.

Spielberger, C. D., Gorsuch, R. L., Lushene, R. E., Vagg, P. R., & Jacobs, G. A. (1983). Manual for the State-Trait Anxiety Inventory. Consulting Psychologists Press.

Tedeschi, R. G., & Calhoun, L. G. (1996). The posttraumatic growth inventory: measuring the positive legacy of trauma. *Journal of Traumatic Stress*, 9, 455–471.

Weiss, D. S., & Marmar, C. R. (1996). The Impact of Event Scale - Revised. In J. Wilson & T.

M. Keane (Eds.), *Assessing psychological trauma and PTSD* (pp. 399-411). Guilford.

## Supplemental File 2 CONSORT Harms 2022 integrated into CONSORT 2010 items checklist of information to include when reporting a randomised trial

CONSORT Harms 2022 integrated into CONSORT 2010 items checklist of information to include when reporting a randomised trial
| Section/Topic | Item No | Checklist item | Reported on page No |
| --- | --- | --- | --- |
| **Title and abstract** |  |  |  |
|  | 1a | Identification as a randomised trial in the title | _____ |
|  | 1b | Structured summary of trial design, methods, results of outcomes of benefits and harms, and conclusions (for specific guidance see CONSORT for abstracts) | _____ |
| **Introduction** |  |  |  |
| Background and objectives | 2a | Scientific background and explanation of rationale | _____ |
|  | 2b | Specific objectives or hypotheses for outcomes benefits and harms | _____ |
| **Methods** |  |  |  |
| Trial design | 3a | Description of trial design (such as parallel, factorial) including allocation ratio | _____ |
|  | 3b | Important changes to methods after trial commencement (such as eligibility criteria), with reasons | _____ |
| Participants | 4a | Eligibility criteria for participants | _____ |
|  | 4b | Settings and locations where the data were collected | _____ |
| Interventions | 5 | The interventions for each group with sufficient details to allow replication, including how and when they were actually administered | _____ |
| Outcomes | 6a | Completely defined pre-specified primary and secondary outcome measures for both benefits and harms, including how and when they were assessed | _____ |
|  | 6b | Any changes to trial outcomes after the trial commenced, with reasons | _____ |
|  | 6c | Describe if and how non-prespecified outcomes of benefits and harms were identified, including any selection criteria, if applicable | _____ |
| Sample size | 7a | How sample size was determined | _____ |
|  | 7b | When applicable, explanation of any interim analyses and stopping guidelines | _____ |
| Randomisation: |  |  |  |
| Sequence generation | 8a | Method used to generate the random allocation sequence | _____ |
|  | 8b | Type of randomisation; details of any restriction (such as blocking and block size) | _____ |
| Allocation concealment mechanism | 9 | Mechanism used to implement the random allocation sequence (such as sequentially numbered containers), describing any steps taken to conceal the sequence until interventions were assigned | _____ |
| Implementation | 10 | Who generated the random allocation sequence, who enrolled participants, and who assigned participants to interventions | _____ |
| Blinding | 11a | If done, who was blinded after assignment to interventions (e.g., participants, care providers, those assessing outcomes of benefits and harms) and how | _____ |
|  | 11b | If relevant, description of the similarity of interventions | _____ |
| Statistical methods | 12a | Statistical methods used to compare groups for primary and secondary outcomes of both benefits and harms | _____ |
|  | 12b | Methods for additional analyses, such as subgroup analyses and adjusted analyses | _____ |
| **Results** |  |  |  |
| Participant flow (a diagram is strongly recommended) | 13a | For each group, the numbers of participants who were randomly assigned, received intended treatment, and were analysed for outcomes of benefits and harms | _____ |
|  | 13b | For each group, losses and exclusions after randomisation, together with reasons | _____ |
| Recruitment | 14a | Dates defining the periods of recruitment and follow-up for outcomes of benefits and harms | _____ |
|  | 14b | Why the trial ended or was stopped | _____ |
| Baseline data | 15 | A table showing baseline demographic and clinical characteristics for each group | _____ |
| Numbers analysed | 16 | For each group, number of participants (denominator) included in each analysis and whether the analysis was by original assigned groups and if any exclusions were made | _____ |
| Outcomes and estimation | 17a | For each primary and secondary outcome of benefits and harms, results for each group, and the estimated effect size and its precision (such as 95% confidence interval) | _____ |
|  | 17a2 | For outcomes omitted from the trial report (benefits and harms), provide rationale for not reporting and indicate where the data on omitted outcomes can be accessed | _____ |
|  | 17b | Presentation of both absolute and relative effect sizes is recommended, for outcomes of benefits and harms | _____ |
|  | 17c | Report zero events if no harms were observed | _____ |
| Ancillary analyses | 18 | Results of any other analyses performed, including subgroup analyses and adjusted analyses, distinguishing pre-specified from exploratory | _____ |
| Harms | 19 | All important harms or unintended effects in each group (for specific guidance see CONSORT for harms) | _____ |
| **Discussion** |  |  |  |
| Limitations | 20 | Trial limitations, addressing sources of potential bias related to the approach to collecting or reporting data on harms, imprecision, and, if relevant, multiplicity or selection of analyses | _____ |
| Generalisability | 21 | Generalisability (external validity, applicability) of the trial findings | _____ |
| Interpretation | 22 | Interpretation consistent with results, balancing benefits and harms, and considering other relevant evidence | _____ |
| **Other information** |  |  |  |
| Registration | 23 | Registration number and name of trial registry | _____ |
| Protocol | 24 | Where the full trial protocol and other relevant documents can be accessed, including additional data on harms | _____ |
| Funding | 25 | Sources of funding and other support (such as supply of drugs), role of funders | _____ |

Note: Adapted from Schulz (2010) to integrate items of CONSORT Harms 2022 (Junqueira 2022) [https://creativecommons.org/licenses/by/2.0/]. CONSORT items 1b, 2b, 6a, 11a, 12a, 13a, 14a, 16a, 17a, 17b, 18, 20 and 24 of were modified to incorporate elements relevant to the reporting of harms. Two new items were added (item 6c and 17a2). Please see the CONSORT Harms 2022 statement for additional details (Junqueira 2022).
We strongly recommend reading the CONSORT 2010 statement (Schulz 2010) in conjunction with the CONSORT Harms 2022 statement (Junqueira 2022) for important clarifications on all the items. If relevant, we also recommend reading CONSORT extensions for cluster randomised trials, non-inferiority and equivalence trials, non-pharmacological treatments, adaptive designs, pilot and feasibility studies, multi arm trials, cross-over and pragmatic trials. Additional extensions are forthcoming: for those and for up-to-date references relevant to this checklist, see EQUATOR Network.

## Supplementary File 3 Incremental Effects

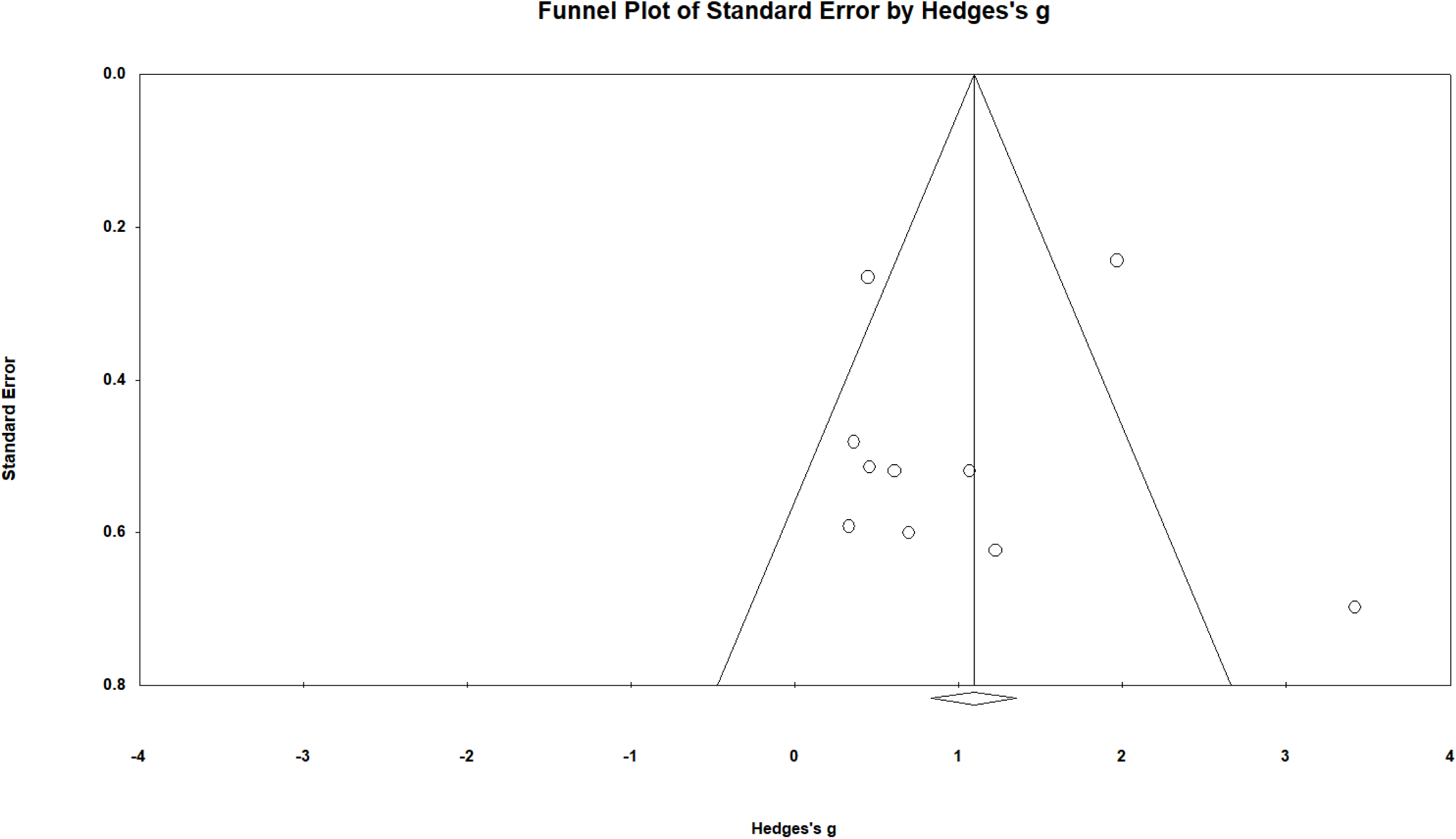

## References

Abdallah, C. G., Roache, J. D., Gueorguieva, R., Averill, L. A., Young-McCaughan, S., Shiroma, P. R., Purohit, P., Brundige, A., Murff, W., Ahn, K. H., Sherif, M. A., Baltutis, E. J., Ranganathan, M., D’Souza, D., Martini, B., Southwick, S. M., Petrakis, I. L., Burson, R. R., Guthmiller, K. B., … Krystal, J. H. (2022). Dose-related effects of ketamine for antidepressant-resistant symptoms of posttraumatic stress disorder in veterans and active duty military: a double-blind, randomized, placebo-controlled multi-center clinical trial. Neuropsychopharmacology, 47, 1574–1581. 10.1038/s41386-022-01266-9

Amoroso, T., & Workman, M. (2016). Treating posttraumatic stress disorder with MDMA-assisted psychotherapy: A preliminary meta-analysis and comparison to prolonged exposure therapy. Journal of Psychopharmacology, 30(7), 595–600. 10.1177/0269881116642542

Armenian, P., & Rodda, L. N. (2022). Two Decades of Ecstasy: Shifting Demographic Trends in Decedents Using MDMA. Journal of Analytical Toxicology, 46, 163–169. 10.1093/jat/bkaa193

Automeris LLC. (2024). Web Plot Digitizer. Automeris.Io. https://automeris.io/WebPlotDigitizer.html

Borenstein, M., Hedges, L., Higgins, J., & Rothstein, H. (2013). Comprehensive Meta-Analysis. Englewood, NJ: Bisostat.

Borgogna, N. C., Owen, T., & Aita, S. L. (2024). The absurdity of the latent disease model in mental health: 10,130,814 ways to have a DSM-5-TR psychological disorder. Journal of Mental Health, 33, 451–459. 10.1080/09638237.2023.2278107

Borgogna, N. C., Owen, T., Petrovitch, D., Vaughn, J., Johnson, D. A. L., Pagano, L. A., Aita, S. L., & Hill, B. D. (2025). Incremental efficacy systematic review and meta-analysis of psilocybin-for-depression RCTs. Psychopharmacology, 242, 2139–2157. 10.1007/s00213-025-06788-w

Borgogna, N. C., Owen, T., Vaughn, J., Johnson, D. A. L., Aita, S. L., & Hill, B. D. (2024). So How Special is Special K? A Systematic Review and Meta-Analysis of Ketamine for PTSD RCTs. European Journal of Psychotraumatology, 15, 2299124.

Brewerton, T. D., Wang, J. B., Lafrance, A., Pamplin, C., Mithoefer, M., Yazar-Klosinki, B., Emerson, A., & Doblin, R. (2022). MDMA-assisted therapy significantly reduces eating disorder symptoms in a randomized placebo-controlled trial of adults with severe PTSD. Journal of Psychiatric Research, 149, 128–135. 10.1016/j.jpsychires.2022.03.008

Ching, T. H. W. (2020). Intersectional insights from an MDMA-assisted psychotherapy training trial: An open letter to racial/ethnic and sexual/gender minorities. Journal of Psychedelic Studies, 4, 61–68. 10.1556/2054.2019.017

Climko, R. P., Roehrich, H., Sweeney, D. R., & Al-Razi, J. (1986). Ecstacy: a review of MDMA and MDA. International Journal of Psychiatry in Medicine, 16, 359–372. 10.2190/dcrp-u22m-aumd-d84h

Cohen, J. (1988). Statistical power analysis for the behavioral sciences (2nd ed.). Lawrence Erlbaum.

Colcott, J., Guerin, A. A., Carter, O., Meikle, S., & Bedi, G. (2025). Side-effects of mdma-assisted psychotherapy: a systematic review and meta-analysis. Neuropsychopharmacology, 49, 1208–1226. 10.1038/S41386-024-01865-8;SUBJMETA

Dalgleish, T., Black, M., Johnston, D., & Bevan, A. (2020). Transdiagnostic approaches to mental health problems: Current status and future directions. Journal of Consulting and Clinical Psychology, 88, 195. 10.1037/CCP0000482

Danforth, A. L., Grob, C. S., Struble, C., Feduccia, A. A., Walker, N., Jerome, L., Yazar-Klosinski, B., & Emerson, A. (2018). Reduction in social anxiety after MDMA-assisted psychotherapy with autistic adults: a randomized, double-blind, placebo-controlled pilot study. Psychopharmacology, 235, 3137–3148. 10.1007/S00213-018-5010-9

De Giorgi, R., & Ede, R. (2024). Psilocybin for depression. BMJ, 385, q798. 10.1136/BMJ.Q798

Deacon, B. J. (2013). The biomedical model of mental disorder: A critical analysis of its validity, utility, and effects on psychotherapy research. Clinical Psychology Review, 33, 846–861. 10.1016/J.CPR.2012.09.007

Dindo, L., Van Liew, J. R., & Arch, J. J. (2017). Acceptance and Commitment Therapy: A Transdiagnostic Behavioral Intervention for Mental Health and Medical Conditions. Neurotherapeutics, 14, 546–553. 10.1007/S13311-017-0521-3

Egger, M., Smith, G. D., Schneider, M., & Minder, C. (1997). Bias in meta-analysis detected by a simple, graphical test. BMJ, 315, 629–634. 10.1136/bmj.315.7109.629

Fadiman, J., & Korb, S. (2019). Might Microdosing Psychedelics Be Safe and Beneficial? An Initial Exploration. Journal of Psychoactive Drugs, 51, 118–122. 10.1080/02791072.2019.1593561

Feduccia, A. A., Jerome, L., Mithoefer, M. C., & Holland, J. (2021). Discontinuation of medications classified as reuptake inhibitors affects treatment response of MDMA-assisted psychotherapy. Psychopharmacology, 238, 581–588. 10.1007/s00213-020-05710-w

Ferguson, C. J. (2009). An effect size primer: A guide for clinicians and researchers. Professional Psychology: Research and Practice, 40, 532–538. 10.1037/a0015808

Gorman, I., Belser, A. B., Jerome, L., Hennigan, C., Shechet, B., Hamilton, S., Yazar-Klosinski, B., Emerson, A., & Feduccia, A. A. (2020). Posttraumatic Growth After MDMA-Assisted Psychotherapy for Posttraumatic Stress Disorder. Journal of Traumatic Stress, 33, 161–170. 10.1002/jts.22479

Harrison, T. R., Faber, S. C., Zare, M., Fontaine, M., & Williams, M. T. (2025). Wolves Among Sheep: Sexual Violations in Psychedelic-Assisted Therapy. American Journal of Bioethics, 25, 40–55. 10.1080/15265161.2024.2433423

Hofmann, S. G., & Hayes, S. C. (2019). The Future of Intervention Science: Process-Based Therapy: Clinical Psychological Science, 7, 37–50. 10.1177/2167702618772296

Insel, T. R., Cuthbert, B., Garvey, M., Heinssen, R., Pine, D. S., Quinn, K., Sanislow, C., & Wang, P. (2010). Research Domain Criteria (RDoC): Toward a New Classification Framework for Research on Mental Disorders. American Journal of Psychiatry, 167, 748–751. 10.1176/APPI.AJP.2010.09091379

Jelovac, A., McCaffrey, C., Terao, M., Shanahan, E., Whooley, E., McDonagh, K., McDonogh, S., Loughran, O., Shackleton, E., Igoe, A., Thompson, S., Mohamed, E., Nguyen, D., O’Neill, C., Walsh, C., & McLoughlin, D. M. (2025). Serial Ketamine Infusions as Adjunctive Therapy to Inpatient Care for Depression: The KARMA-Dep 2 Randomized Clinical Trial. JAMA Psychiatry, 82, 1216–1224. 10.1001/jamapsychiatry.2025.3019

Jerome, L., Feduccia, A. A., Wang, J. B., Hamilton, S., Yazar-Klosinski, B., Emerson, A., Mithoefer, M. C., & Doblin, R. (2020). Long-term follow-up outcomes of MDMA-assisted psychotherapy for treatment of PTSD: a longitudinal pooled analysis of six phase 2 trials. Psychopharmacology, 237, 2485–2497. 10.1007/s00213-020-05548-2

Junqueira, D. R., Zorzela, L., Golder, S., Loke, Y., Gagnier, J. J., Julious, S. A., Li, T., Mayo-Wilson, E., Pham, B., Phillips, R., Santaguida, P., Scherer, R. W., Gøtzsche, P. C., Moher, D., Ioannidis, J. P. A., & Vohra, S. (2023). CONSORT Harms 2022 statement, explanation, and elaboration: updated guideline for the reporting of harms in randomised trials. BMJ, 381. 10.1136/BMJ-2022-073725

Krystal, J. H., Rosenheck, R. A., Cramer, J. A., Vessicchio, J. C., Jones, K. M., Vertrees, J. E., Horney, R. A., Huang, G. D., & Stock, C. (2011). Adjunctive Risperidone Treatment for Antidepressant-Resistant Symptoms of Chronic Military Service–Related PTSD: A Randomized Trial. JAMA, 306, 493–502. 10.1001/JAMA.2011.1080

Lamotte, S. (2022, November 3). Severe depression eased by single dose of synthetic ‘magic mushroom.’ CNN. https://www.cnn.com/2022/11/02/health/psilocybin-magic-mushroom-depression-wellness/index.html

Mitchell, J. M., Bogenschutz, M., Lilienstein, A., Harrison, C., Kleiman, S., Parker-Guilbert, K., Ot’alora G, M., Garas, W., Paleos, C., Gorman, I., Nicholas, C., Mithoefer, M., Carlin, S., Poulter, B., Mithoefer, A., Quevedo, S., Wells, G., Klaire, S. S., van der Kolk, B., … Doblin, R. (2021). MDMA-assisted therapy for severe PTSD: a randomized, double-blind, placebo-controlled phase 3 study. Nature Medicine, 27, 1025–1033. 10.1038/s41591-021-01336-3

Mitchell, J. M., Ot’alora G, M., van der Kolk, B., Shannon, S., Bogenschutz, M., Gelfand, Y., Paleos, C., Nicholas, C. R., Quevedo, S., Balliett, B., Hamilton, S., Mithoefer, M., Kleiman, S., Parker-Guilbert, K., Tzarfaty, K., Harrison, C., de Boer, A., Doblin, R., & Yazar-Klosinski, B. (2023). MDMA-assisted therapy for moderate to severe PTSD: a randomized, placebo-controlled phase 3 trial. Nature Medicine, 29, 2473–2480. 10.1038/s41591-023-02565-4

Mithoefer, M. C., Feduccia, A. A., Jerome, L., Mithoefer, A., Wagner, M., Walsh, Z., Hamilton, S., Yazar-Klosinski, B., Emerson, A., & Doblin, R. (2019). MDMA-assisted psychotherapy for treatment of PTSD: study design and rationale for phase 3 trials based on pooled analysis of six phase 2 randomized controlled trials. Psychopharmacology, 236, 2735–2745. 10.1007/s00213-019-05249-5

Mithoefer, M. C., Mithoefer, A. T., Feduccia, A. A., Jerome, L., Wagner, M., Wymer, J., Holland, J., Hamilton, S., Yazar-Klosinski, B., Emerson, A., & Doblin, R. (2018). 3,4-methylenedioxymethamphetamine (MDMA)-assisted psychotherapy for post-traumatic stress disorder in military veterans, firefighters, and police officers: a randomised, double-blind, dose-response, phase 2 clinical trial. The Lancet Psychiatry, 5, 486–497. 10.1016/S2215-0366(18)30135-4

Mithoefer, M. C., Wagner, M. T., Mithoefer, A. T., Jerome, L., & Doblin, R. (2011). The safety and efficacy of ±3,4-methylenedioxymethamphetamine- assisted psychotherapy in subjects with chronic, treatment-resistant posttraumatic stress disorder: The first randomized controlled pilot study. Journal of Psychopharmacology, 25, 439–452. 10.1177/0269881110378371

National Institute of Mental Health. (2022). Mental Illness Statistics. U.S. Department of Health and Human Services, National Institutes of Health. https://www.nimh.nih.gov/health/statistics/mental-illness

Nicholas, C. R., Wang, J. B., Coker, A., Mitchell, J. M., Klaire, S. S., Yazar-Klosinski, B., Emerson, A., Brown, R. T., & Doblin, R. (2022). The effects of MDMA-assisted therapy on alcohol and substance use in a phase 3 trial for treatment of severe PTSD. Drug and Alcohol Dependence, 233. 10.1016/j.drugalcdep.2022.109356

Oehen, P., Traber, R., Widmer, V., & Schnyder, U. (2013). A randomized, controlled pilot study of MDMA (± 3,4-Methylenedioxymethamphetamine)-assisted psychotherapy for treatment of resistant, chronic Post-Traumatic Stress Disorder (PTSD). Journal of Psychopharmacology, 27, 40–52. 10.1177/0269881112464827

Ot’alora G, M., Grigsby, J., Poulter, B., Van Derveer, J. W., Giron, S. G., Jerome, L., Feduccia, A. A., Hamilton, S., Yazar-Klosinski, B., Emerson, A., Mithoefer, M. C., & Doblin, R. (2018). 3,4-Methylenedioxymethamphetamine-assisted psychotherapy for treatment of chronic posttraumatic stress disorder: A randomized phase 2 controlled trial. Journal of Psychopharmacology, 32, 1295–1307. 10.1177/0269881118806297

Pantoni, M. M., Kim, J. L., Van Alstyne, K. R., & Anagnostaras, S. G. (2022). MDMA and memory, addiction, and depression: dose-effect analysis. Psychopharmacology, 239(3), 935–949. 10.1007/s00213-022-06086-9

Parrott, A. C. (2007). The psychotherapeutic potential of MDMA (3,4-methylenedioxymethamphetamine): an evidence-based review. Psychopharmacology, 191, 181–193. 10.1007/s00213-007-0703-5

Passie, T. (2018). The early use of MDMA (‘Ecstasy’) in psychotherapy (1977–1985). *Drug Science*, Policy and Law, 4. 10.1177/2050324518767442

Pizer, J. H., Aita, S. L., Myers, M. A., Hawley, N. A., Ikonomou, V. C., Brasil, K. M., Hernandez, K. A., Pettway, E. C., Owen, T., Borgogna, N. C., Smitherman, T. A., & Hill, B. D. (2024). Neuropsychological Function in Migraine Headaches. Neurology, 102(4), e208109. 10.1212/WNL.0000000000208109/SUPPL_FILE/SUPPLEMENTARY_DATA1.PDF

Ponte, L., Jerome, L., Hamilton, S., Mithoefer, M. C., Yazar-Klosinski, B. B., Vermetten, E., & Feduccia, A. A. (2021). Sleep Quality Improvements After MDMA-Assisted Psychotherapy for the Treatment of Posttraumatic Stress Disorder. Journal of Traumatic Stress, 34, 851–863. 10.1002/jts.22696

Rhee, T. G., Davoudian, P. A., Sanacora, G., & Wilkinson, S. T. (2023). Psychedelic renaissance: Revitalized potential therapies for psychiatric disorders. Drug Discovery Today, 28, 103818. 10.1016/J.DRUDIS.2023.103818

Schenberg, E. E. (2018). Psychedelic-assisted psychotherapy: A paradigm shift in psychiatric research and development. Frontiers in Pharmacology, 9, 323606. 10.3389/FPHAR.2018.00733/BIBTEX

Schifano, F. (2004). A bitter pill. Overview of ecstasy (MDMA, MDA) related fatalities. Psychopharmacology, 173, 242–248. 10.1007/s00213-003-1730-5

Shahrour, G., Sohail, K., Elrais, S., Khan, M. H., Javeid, J., Samdani, K., Mansoor, H., Hussain, S. I., Sharma, D., Ehsan, M., & Nashwan, A. J. (2024). MDMA-assisted psychotherapy for the treatment of PTSD: A systematic review and meta-analysis of randomized controlled trials (RCTs). Neuropsychopharmacology Reports, 44, 672–681. 10.1002/npr2.12485

Sicignano, D. J., Kurschner, R., Weisman, N., Sedensky, A., Hernandez, A. V., & White, C. M. (2024). The Impact of Ketamine for Treatment of Post-Traumatic Stress Disorder: A Systematic Review With Meta-Analyses. Annals of Pharmacotherapy, 58, 669–677. 10.1177/10600280231199666

Siegel, J. S., Daily, J. E., Perry, D. A., & Nicol, G. E. (2023). Psychedelic Drug Legislative Reform and Legalization in the US. JAMA Psychiatry, 80, 77–83. 10.1001/jamapsychiatry.2022.4101

Smith, K. W., Sicignano, D. J., Hernandez, A. V., & White, C. M. (2022). MDMA-Assisted Psychotherapy for Treatment of Posttraumatic Stress Disorder: A Systematic Review With Meta-Analysis. Journal of Clinical Pharmacology, 62, 463–471. 10.1002/jcph.1995

Taillefer de Laportalière, T., Jullien, A., Yrondi, A., Cestac, P., & Montastruc, F. (2023). Reporting of harms in clinical trials of esketamine in depression: a systematic review. Psychological Medicine, 1–11. 10.1017/S0033291723001058

The White House. (2022). Reducing the Economic Burden of Unmet Mental Health Needs. The White House. https://www.whitehouse.gov/cea/written-materials/2022/05/31/reducing-the-economic-burden-of-unmet-mental-health-needs/

van der Kolk, B. A., Wang, J. B., Yehuda, R., Bedrosian, L., Coker, A. R., Harrison, C., Mithoefer, M., Yazar-Klosinki, B., Emerson, A., & Doblin, R. (2024). Effects of MDMA- assisted therapy for PTSD on self-experience. PloS One, 19. 10.1371/journal.pone.0295926

von Rotz, R., Schindowski, E. M., Jungwirth, J., Schuldt, A., Rieser, N. M., Zahoranszky, K., Seifritz, E., Nowak, A., Nowak, P., Jäncke, L., Preller, K. H., & Vollenweider, F. X. (2023). Single-dose psilocybin-assisted therapy in major depressive disorder: A placebo-controlled, double-blind, randomised clinical trial. The Lancet, 56, 101809. 10.1016/j.eclinm.2022.101809

Wolfson, P. E., Andries, J., Feduccia, A. A., Jerome, L., Wang, J. B., Williams, E., Carlin, S. C., Sola, E., Hamilton, S., Yazar-Klosinski, B., Emerson, A., Mithoefer, M. C., & Doblin, R. (2020). MDMA-assisted psychotherapy for treatment of anxiety and other psychological distress related to life-threatening illnesses: a randomized pilot study. Scientific Reports, 10(1). 10.1038/s41598-020-75706-1

